# Role of treatments for diabetes and hyperlipidaemia in risk and mortality of primary and secondary brain tumours

**DOI:** 10.1101/2020.09.20.20198325

**Authors:** Jamie W Robinson, Richard M Martin, Mio Ozawa, Martha M C Elwenspoek, Maria Theresa Redaniel, Kathreena M Kurian, Yoav Ben-Shlomo

**Affiliations:** MRC Integrative Epidemiology Unit, University of Bristol, Bristol, BS8 2BN, UK; Department of Population Health Sciences, University of Bristol, Bristol, BS8 1UD, UK; National Institute for Health Research (NIHR) Bristol Biomedical Research Centre, University Hospitals Bristol and University of Bristol, Bristol, UK; Population, Policy and Practice Research and Teaching Department, UCL Great Ormond Street Institute of Child Health, 30 Guilford Street, London, WC1N 1EH; The National Institute for Health Research Applied Research Collaboration West (NIHR ARC West), University Hospitals Bristol NHS Foundation Trust, BS1 2NT Bristol, UK; Brain Tumour Research Centre, Bristol, BS10 5NB, UK

**Keywords:** Primary and secondary brain tumours, fibrates, glitazones, observational epidemiology

## Abstract

**Background:** *In vivo* and observational studies suggest anti-hyperlipidaemic and -diabetic medications (fibrates and glitazones respectively) may have a role in primary prevention and progression of brain tumours by targeting PPAR-α and -γ, respectively.

**Methods:** We conducted a case-control and clinical cohort study within the UK Clinical Practice Research Datalink. We identified adults (aged 18 years+) with primary or secondary brain tumours diagnosed between 2000-2016 prescribed either fibrates or glitazones and identified four controls based on age, sex and drug exposure duration. Multivariable logistic regression analysis estimated an association between drug exposure and brain tumour status. Cox’s survival models were used to look at risk of mortality.

**Results:** 1,916 cases were prescribed a fibrate and 445 cases a glitazone. Our analyses showed little evidence of an association between fibrates and either risk or mortality of brain tumours (adjusted odds ratio for ever exposed PPAR-α 0.98, 95% CI: 0.77, 1.23; adjusted hazard ratio for ever exposed 0.91; 95% CI: 0.76, 1.09). We observed a reduced risk with a per-year increase in exposure duration for glitazones (adjusted odds ratio 0.88, 95% CI: 0.81, 0.96, P=0.002) but no major mortality benefit (adjusted hazard ratio 0.99, 95% CI: 0.80, 1.23).

**Conclusions:** Our results suggest longer duration exposure to glitazones is associated with a reduced risk of primary and secondary brain tumours but no large effect on mortality. We failed to find any strong evidence of a protective effect on risk or mortality for fibrate exposure. Further studies are required for replication and to provide more precise effect estimates.

**Key Points:**

- Repurposing existing drugs that target PPAR-α and -γ receptors may have a role in prevention and progression of brain tumours.
- We observed a reduced risk between duration of glitazones and both primary and secondary brain tumours but no major reduction on mortality
- We observed no major reduction of risk or mortality with fibrates
- Future studies need to be undertaken to ensure replication and obtain more precise estimates

**Importance of Study:** The incidence of brain tumours appears to be increasing with a growing impact on years of life lost. Therapeutic options are limited, either for primary prevention or to prevent mortality of disease once developed. We investigated whether two commonly prescribed families of drugs (fibrates and glitazones) could offer potential drug-repurposing options in reducing brain tumour incidence or progression as informed by previous *in vivo* and observational studies. Our analyses suggest that glitazones are associated with a decreased brain tumour risk. These results can help guide future research into drug re-repurposing for brain tumour treatment.

## Introduction

Brain, other central nervous system (CNS) and intracranial tumours are the 9^th^ most common cancer in the UK with an incidence of 18 per 100,000 individuals in 2016 ^1^. Brain cancer prognosis varies greatly by histological grade. Glioblastoma (WHO grade IV) has an extremely poor outlook, with roughly 5% of patients alive after five years ^2^. Drug companies are increasingly withdrawing from the development of novel drugs due to significant costs and high failure rates, which is especially true for cancer therapeutics ^3^. However, re-purposing existing approved drugs for different diseases than originally designed provides significant advantages over how drugs are traditionally developed saving substantial time and costs involved in conducting new clinical trials.

Nuclear hormone receptors (NHR) have crucial roles in cellular homeostasis and have been implicated in the development of cancer. Peroxisome proliferator-activated receptors (PPARs) are a family of ligand-activated transcription factors that regulate the expression of genes and, as a subtype of NHRs, are involved in the control of proliferation and differentiation of cells ^4-6^. This has marked PPARs as potential candidates in the treatment of cancer, with some studies showing beneficial outcomes in animal models and some early-stage human trials ^7-10^. There are three isoforms of PPARs which are designed as -α, -β/δ and -γ. The genes regulated by these transcription factors are also involved in the transport, metabolism and storage of fatty acids, inflammation and diabetes mellitus ^5,11,12^. PPAR-α and -γ are of considerable clinical significance due to the existence of agonistic compounds that these bind to, namely hypolipidaemic fibrates and thiazolidinediones (glitazones) respectively.

PPAR-α expression has been seen to be enriched within the classical glioblastoma subtype and constituted an independent prognostic marker for improved overall survival ^13^. PPAR-γ is increased in human breast cancer, and ligand activation of this receptor results in a more differentiated and thus less malignant state of the disease ^14^ and reduced growth of colon cancer cells ^15^. Pioglitazone, a PPAR-γ agonist, showed a dose-dependent reduction of glioma tumour invasion in murine glioma models when combined with 6-OH-11-O-hydroxyfenantrene (IIF) ^16^.

Human evidence is limited with a meta-analysis of 17 randomised placebo-controlled trials, and a separate phase II trial, suggesting that PPAR-α agonists may not succeed as anti-cancer agents in general ^17 18^. However, the first of these studies did not look directly at CNS cancer and the second of these studies did not have a large cohort of CNS cancer cases (n = 97). A previous case control study found that diabetic glioblastoma patients were less likely to be treated with a PPAR-γ agonist when compared to a hip fracture control population ^19^, though the study was potentially biased as thiazolidinedione use may be over-represented amongst people with fractures ^20,21^. We have explored whether PPAR-α or -γ agonists are associated with a reduced risk of developing brain tumours or mortality risk using a large population-based database from primary care.

## Methods and Materials

To examine the effects of PPAR-α and -γ exposure, we undertook two nested case-control studies and a case-only clinical cohort study within the UK Clinical Practice Research Datalink (CPRD). CPRD is a primary care database with clinical information on over 11 million people from more than 670 UK GP practices ^22^. The study was approved by the Independent Scientific Advisory Committee (ISAC) for the UK Medicines and Healthcare Products Regulatory Agency (ISAC protocol number: 18_149R ^23^). The data were extracted from CPRD GOLD and linked to ONS death registration data and census data on area deprivation (see below).

### Participants

All participants in the study were 18 years or older and were registered within the CPRD between 1^st^ January 2000 to 1^st^ January 2016, the former being the first year that glitazones were licensed within the UK. Follow-up was stopped when one of the following occurred: death; brain tumour diagnosis; end of registration at a CPRD GP practice; or the end-date of the study. We specified for inclusion that brain tumour patients must have received their diagnosis after registration at a CPRD participating GP practice, due to the possibility that their prescription history may be missing or incomplete.

All participants in the case control study had to be treated with either an anti-hyperlipidaemic or anti-diabetic medication before the end date of the study to reduce the likelihood of confounding by indication. However, those people treated solely with insulin therapy were dropped so as to exclude subjects with type I diabetes. Participants who started on oral medication, but at some point received insulin therapy, were still eligible for inclusion. We did not exclude people on combination therapy (for example, both glitazone plus another anti-diabetic medication) that included the drug of interest; these participants were considered as exposed, if one of the therapies was the drug of interest. Similarly, some people were exposed to both fibrates and glitazones and we considered these exposed for both analyses.

In the cohort study, all brain tumour patients, regardless of fibrate or glitazone drug exposure were included so long as they had a minimum of one year of follow-up of observation prior to censoring.

### Cases and Controls

We defined cases as those patients who were diagnosed with a brain tumour (primary or secondary) and had this diagnosis recorded in the CPRD. We included secondary tumours as fibrates and glitazones may theoretically effect risk of other tumours having metastatic spread to the brain. A list of the read codes and CPRD descriptions of these can be found in **Supplementary Table 1**.

Up to four controls, without diagnosis of a brain tumour and contemporaneously registered within the CPRD, were selected per case using incidence density sampling. Controls were subject to the same selection criteria as cases and were strata matched by age group (<20, 20-29, 30-39, 40-49, 50-59, 60-69, 70-79 and ≥80) and sex. To address time-window bias ^24^, controls were required to have the same retrospective duration of potential exposure (any drug treatment for either hyperlipidaemia or diabetes) within the CPRD as cases, based on the case index date plus or minus six months. For example, if a case had 6.3 years of retrospective drug history from their index date in 2013, then only controls who also had between 5.8 and 6.8 years of drug exposure over the same secular time period were sampled. This meant that both cases and controls have the same potential for recorded exposure to the drugs of interest.

### Exposures

Our drug exposures were PPAR-α and -γ agonists for the treatment of hyperlipidaemia and type II diabetes as compared to another drug treatment (unexposed group), respectively, with product codes used to determine this given in **Supplementary Tables 2** and **3**.

We created two exposure variables. Firstly, a binary variable to indicate ever exposed to either drug as compared to another potential drug for the management of diabetes or hyperlipidaemia. Secondly, the total time the participant had been exposed to one of the drugs before censoring, which we categorised into an ordinal variable representing years of exposure for use in the logistic regression model. This was calculated by summing each uninterrupted prescription duration for each person. We defined an interruption as a break of at least 90 days between prescriptions, which started a new uninterrupted duration. The longest of these prescription durations were used to determine exposure duration. As we did not know whether any latency period is required to have a physiological effect, we used this duration variable to investigate a potential dose-response relationship. Exposure duration was categorised into yearly categories: 0 or exposure ≤ 1 year, >1 and ≤ 2 years, >2 and ≤ 3 years, >3 and ≤ 4 years, >4 and ≤ 5 years, >5 and ≤ 6 years, >6 and ≤ 7 years and > 7 years exposure due to small numbers of people reaching this length of exposure.

### Potential Confounders

We considered the following variables as potential confounders that might influence both risk of developing or dying from a brain tumour as well as potential influencing the choice of drug agent that a doctor might prescribe; these were age, sex and socioeconomic status (SES). We used an ecological proxy measure, the Index of Multiple Deprivation 2015 (IMD) ^25^, to proxy for socioeconomic status. This is a commonly used measure in the United Kingdom that uses census data on a wide variety of economic and health factors to derive a postcode-based deprivation score so that that a higher score indicates less deprivation. These were grouped into five equal sized groups after sorting (quintiles).

In the analyses for diabetic medications, we felt it was important to adjust for the severity or degree of diabetic control as a potential confounder. We did this by using, where available, HbA1c levels. Units were converted and standardised to the International Federation of Clinical Chemistry (IFCC) units in mmols/mol. We created an ordinal variable with three levels: 1, indicating well controlled diabetes for levels ≤ 58 mmols/mol; 2, indicating sub-optimally controlled diabetes for levels > 58 mmols/mol and ≤ 75 mmols/mol; and 3, indicating poorly controlled diabetes for levels > 75 mmols/mol. If no measures of the participant’s HbA1c levels were identified in the database, we assumed they had mild or well controlled diabetes and assigned these participants into the lowest ordinal level based on the assumption that the clinician did not feel it necessary to do the blood test. However, we also conducted sensitivity analyses to test this assumption (see below).

For the cohort analysis we assumed patients with more comorbidities would be more likely to die and this may also influence choice of medication. We therefore derived the Charlson comorbidity index score ^26^ in the cohort analysis and included this as a covariate. The list of read codes used was taken from Khan, *et al*, ^27^ and result in an ordinal score. We calibrated this to 4 units (half the interquartile range) so that the model coefficient is for a 4-unit change in Charlson comorbidity index score.

### Statistical Methods

The fibrate and glitazone case-control analyses were conducted using logistic regression to compute odds ratios (OR) and 95% confidence intervals (CI) for exposure status to fibrate and glitazone drugs and exposure duration of the two types of drugs and case control status. We ran unadjusted and multivariable models adjusting for age, sex, quintile IMD score, drug treatment duration and HbA1c for the glitazone study. We chose to run an unmatched logistic rather than conditional logistic model as this may introduce bias but we treated matching variables as covariates ^28^. Dose exposure was analysed both as a continuous ordinal variable but also as a “dummy” variable so we could check for any evidence of non-linearity in the pattern of the odds ratios.

The cohort analysis consisted of both unadjusted Kaplan-Meier graphs and adjusted Cox proportional hazards models with estimated hazard ratios (HR) and 95% confidence intervals. The proportional hazards assumption was tested by examination of the scaled Schoenfeld residuals.

### Sensitivity Analyses

We repeated the analyses with primary and secondary brain tumours, rather than the combined group, to see if there was any evidence of heterogeneity of effect. We also investigated our assumption that missing HbA1c levels should be allocated to the well-controlled group. We undertook multiple imputation to predict missing HbA1c levels based on case control status, age, IMD, sex, drug treatment duration, PPAR-γ ever exposure and number of consultations, defined as each day the patient had at least one in-person consultation. We generated 55 datasets, roughly equal to the missingness of the data, and these were then combined using Rubin’s rules. We also repeated the analyses on the complete case subset so that any subjects with missing HbA1c levels were dropped.

### Libraries and Code

We used Python and the Pandas ^29^ library to clean the data and construct the summary statistics for the models. The logistic regression and Kaplan-Meier curves were conducted in R using the Survival library ^30^. The Survminer library ^31^ in R was also used for the Cox proportional hazards model. MICE ^32^ was used to impute missing HbA1c levels.

## Results

After data extraction, cleaning and linkage, the study populations consisted of 9,741 participants with 129,356 person years in the fibrate study and 2,400 participants with 30,871 person years of follow up in the glitazone study (see flow charts for participant selection in **Supplementary Figures 1-3**).

Cases and controls had similar distributions for sex, age group and quintile IMD score (see **Table 1**). Cases appeared to have similar probability of exposure to fibrates but less exposure to glitazones compared to controls. This was supported by the multivariable models (**Table 2**). For fibrates there was little evidence that ever exposed (adjusted OR (aOR)=0.98; 95% CI: 0.77 to 1.23; P=0.84) or per year increase in duration of exposure (aOR=0.97; 95% CI: 0.91 to 1.03; P=0.34) was associated with risk. There was also little evidence of a dose-response relationship between fibrate exposure and brain tumour risk either as a continuous or dummy variable (**see Supplementary Table 4**).

**Table 1.**
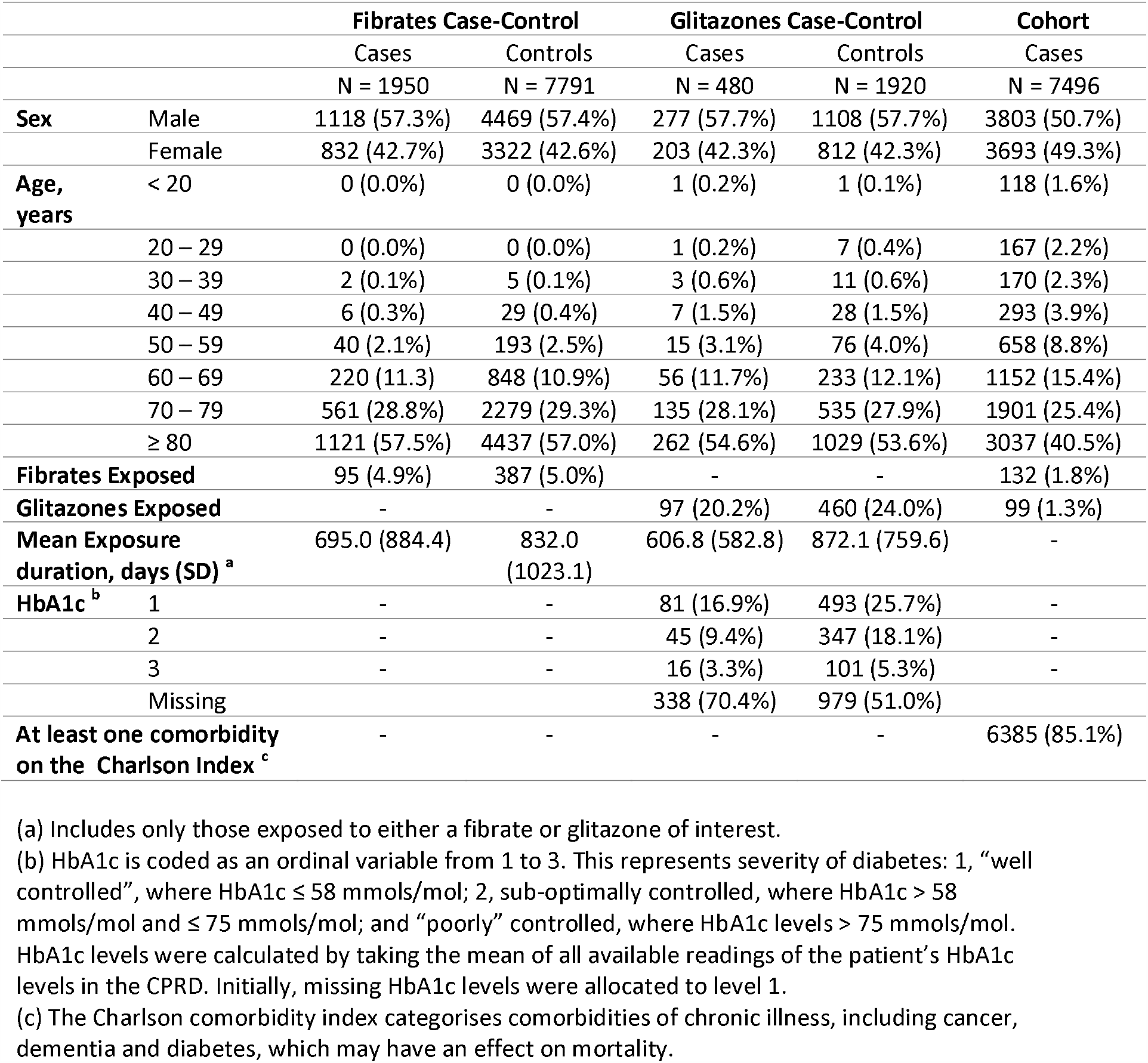
Patient characters in each of the three datasets we used.

**Table 2.**
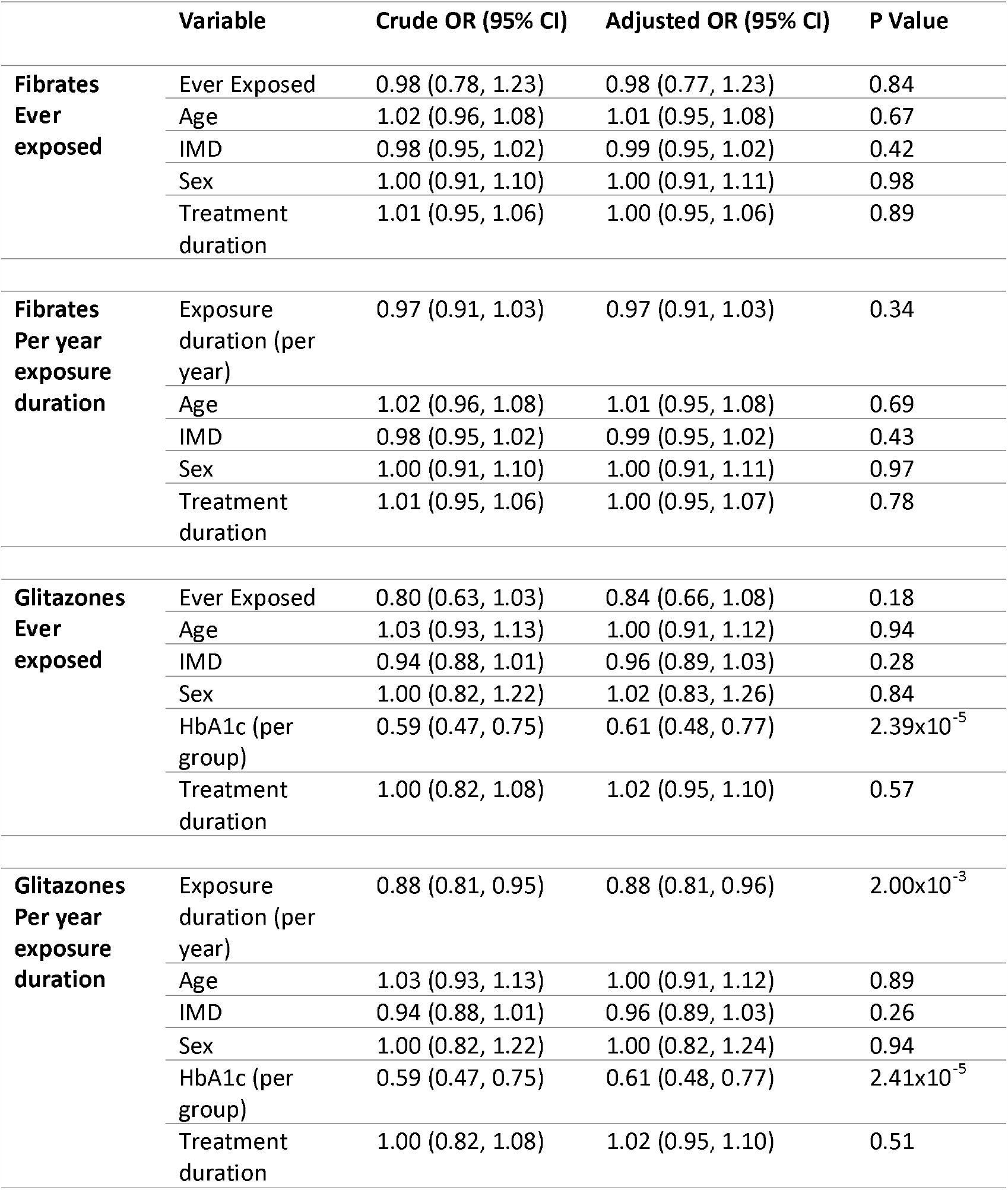
Results from the fibrate and glitazone logistic regressions.

**Table 3.** Results from the fibrates and glitazones Cox proportional hazards model.

|  | **Variable** | **Adjusted HZ (95% CI)** | **P Value** |
| --- | --- | --- | --- |
| **Fibrates**<br>**Ever exposed** | Ever Exposed | 0.90 (0.75, 1.07) | 0.24 |
| | Age | 1.36 (1.29, 1.43) | $< 2.00 \times 10^{-16}$ |
|  | IMD | 1.02 (0.99, 1.05) | 0.28 |
|  | Sex | 0.92 (0.84, 1.00) | 0.05 |
| | Charlson | 1.14 (1.11, 1.17) | $< 2.00 \times 10^{-16}$ |
| **Glitazones**<br>**Ever Exposed** | Ever Exposed | 0.97 (0.78, 1.20) | 0.77 |
| | Age | 1.23 (1.16, 1.31) | $4.79 \times 10^{-11}$ |
|  | IMD | 0.99 (0.94, 1.04) | 0.65 |
|  | Sex | 0.96 (0.84, 1.11) | 0.60 |
| | Charlson | 1.14 (1.09, 1.20) | $1.71 \times 10^{-7}$ |

For glitazones, the association with ever exposure was consistent with chance (aOR=0.84; 95% CI: 0.66, 1.08; P=0.18) but there was an inverse association with duration of exposure (aOR per year = 0.88; 95% CI: 0.81, 0.96; P=2.00×10^−3^). Analysis by duration period was consistent with an effect only being observed after 4 years (**see Supplementary Table 5**) as all the odds ratio were less than one, however the confidence intervals for the shorter duration periods were sufficiently wide as to not exclude any effect for earlier periods. These results were almost identical when we imputed the missing HbA1c values (aOR=0.88, 95% CI: 0.82, 0.94, P=1×10^−3^) or if we did not adjust for HbA1c (**see Supplementary Table 6**). However, in the complete case analysis the effect was attenuated to the null (aOR 0.96, 95% CI 0.85, 1.08, P>0.05). We noted that higher HbA1C levels were also associated with a reduced risk (aOR=0.61; 95% CI: 0.48, 0.77; P=2.41×10^−5^). However, in both the imputed and complete case analysis there was no association between HbA1c and case control status (aOR=0.99, 95% CI: 0.82 to 1.20 and aOR=0.94, 95% CI: 0.71, 1.24) (see **Supplementary Table 6**).

The repeat analysis looking at primary and secondary tumours alone found similar results (aOR=0.89 and 0.87 respectively) (**see supplementary table 7**). We were underpowered to examine whether any effects may have differed by type of specific cancers e.g. lung, breast that were the primary source for the secondary brain tumours.

In the cohort study, there was little evidence that exposure to fibrates improved survival rates when compared to the non-exposed population (unadjusted exposed median survival time = 3.26 months; 95% CI: 2.60, 4.15 months versus unadjusted unexposed median survival time = 3.49 months; 95% CI: 3.26, 3.75 months) (**Figure 1**). There was also little evidence to suggest that glitazones were associated with survival rates (unadjusted exposed median survival time = 3.29 months; 95% CI: 2.86, 5.43 months versus unadjusted unexposed median survival time = 4.64; 95% CI: 3.92, 5.56 months) (**Figure 2**). There was little evidence of an association between fibrate exposure and risk of death (adjusted hazard ratio, aHR=0.91; 95% CI: 0.76, 1.09; P=0.30). There was also little evidence of a decreased risk of death associated with glitazones (aHR=0.99; 95% CI: 0.80, 1.23; P=0.95).

**Figure 1.**
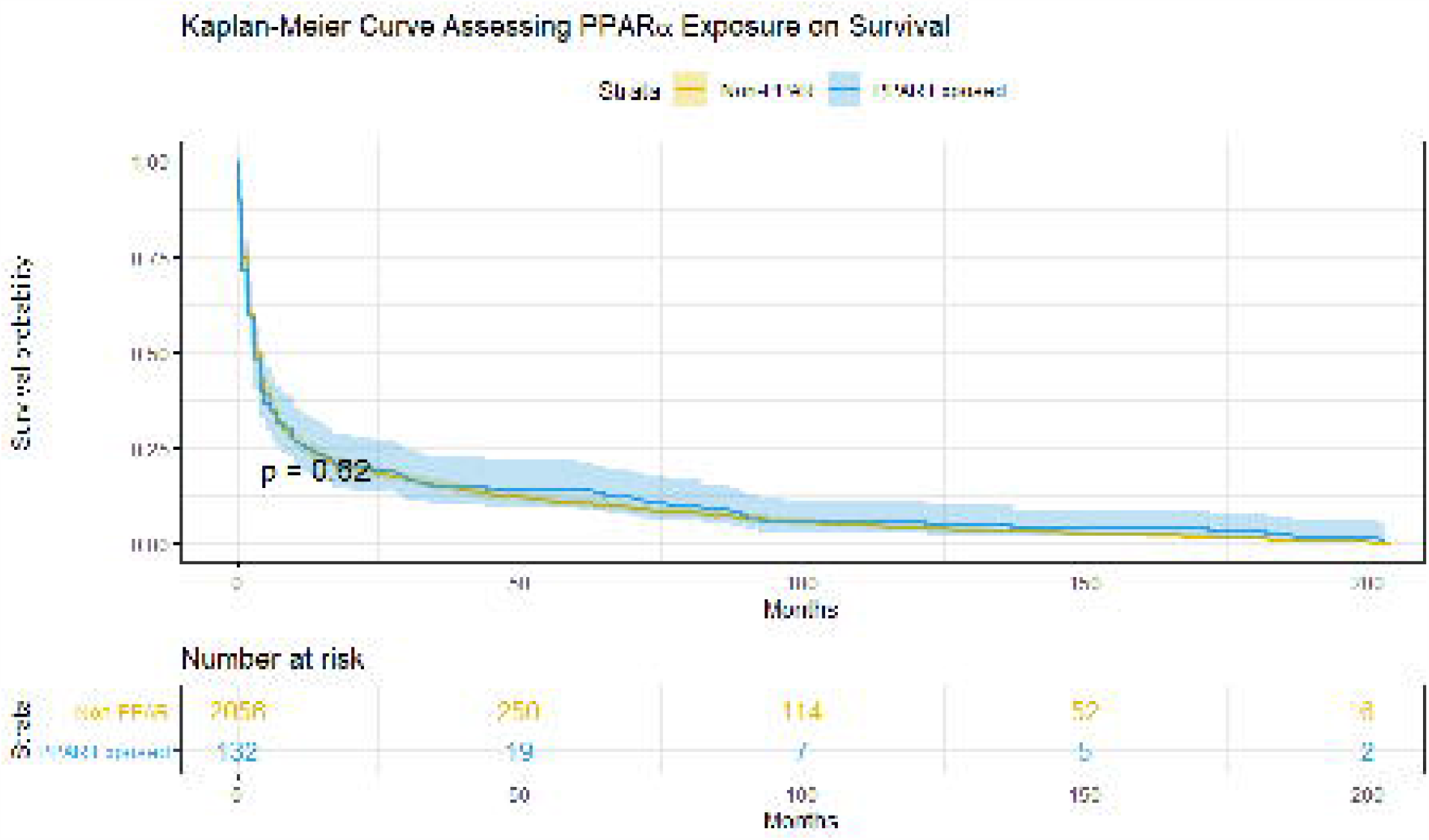
Kaplan-Meier curve for cohort analysis investigating exposure to fibrates (PPAR-α agonist).

**Figure 2.**
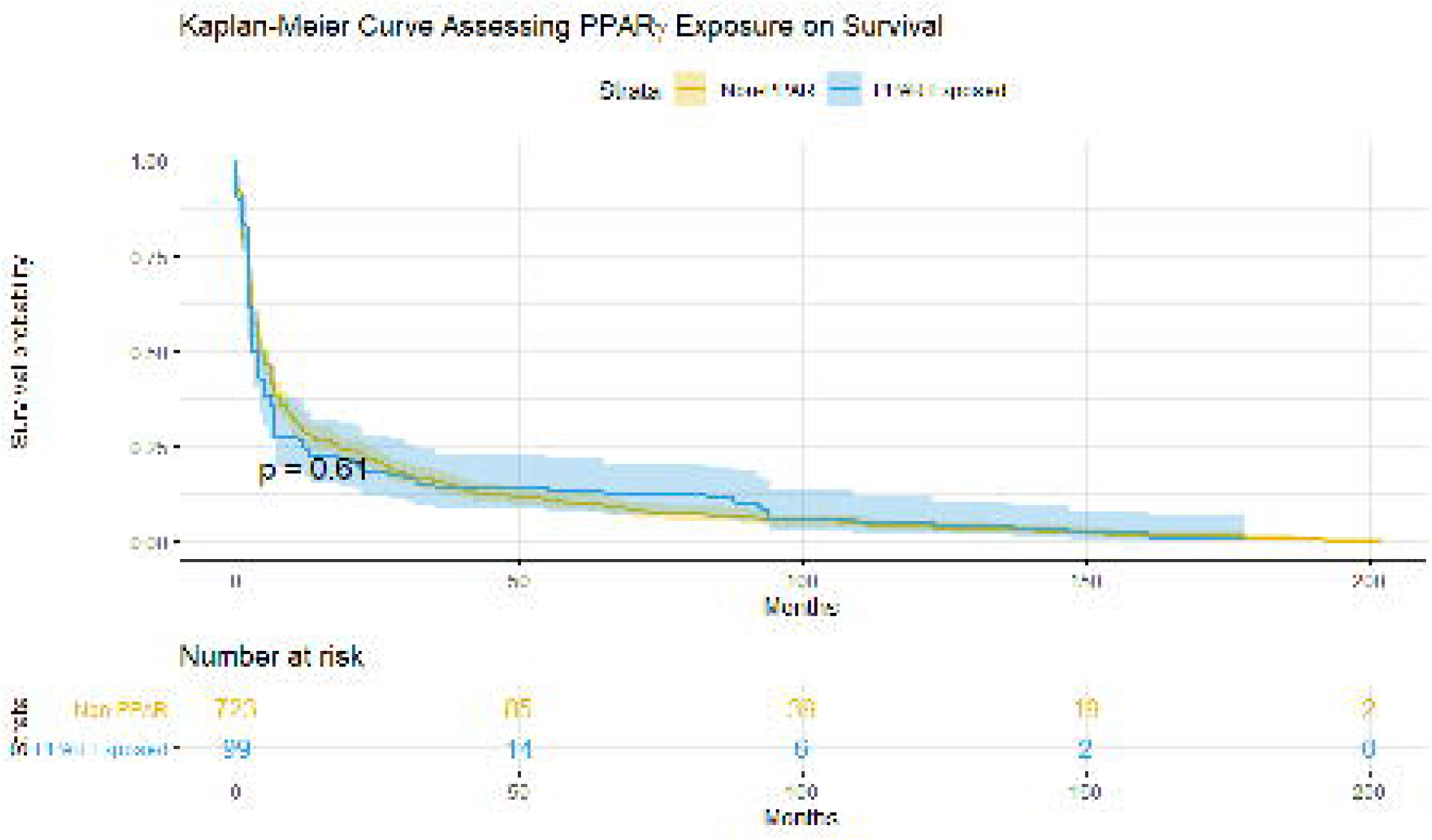
Kaplan-Meier curve for cohort analysis investigating exposure to glitazones (PPAR-γ agonist).

## Discussion

To the best of our knowledge, this is the first pharmacoepidemiological study of the effects of fibrate and glitazone prescription on risk and progression of brain tumours. We found little evidence that fibrates were protective against brain tumour risk or mortality, in contrast to some previous literature ^33^. We did observe that ever exposure to glitazones showed an inverse effect with risk which seemed most marked for exposure duration of 4 or more years. This observation was consistent when we used multiple imputation for missing HbA1C levels rather than assume this reflected good glycaemic control. However, the effect was attenuated in the complete case analysis. The latter however resulted in far fewer cases and reduced precision in our estimated risk (95% CI: 0.85, 1.08) which were still consistent with our original observation. It is also more prone to collider bias ^34^, whereby controlling for a common effect of both the exposure (glitazone prescription) and the outcome (brain tumour) distorts the association between the two. This would introduce an artefactual association between treatment and brain tumours if glitazones and brain tumours are associated with having a HbA1c test. This is possible as in the UK, glitazones are recommended as second line therapy for diabetes ^35^ and hence given to less easy to control diabetics who are more likely to require greater monitoring. Brain tumour risk may also be associated with testing if risk factors or confounders are associated with probability of testing.

If our observed association with glitazones is causal, then there may be a potential protective effect for glitazones, although this observation requires replication with ideally a larger sample size so one can test if there is a genuine threshold effect with duration. Similar effects were seen when considering only primary or only secondary brain tumours but due to limited power we could not test whether there were differences by type of cancer for those subjects with secondary brain tumours. In the case of fibrates, however, no such effect was observed suggesting that they do not alter brain tumour risk or mortality.

This study uses CPRD a large and well-established database which has been validated by numerous sources ^36-38^. It should be free of selection bias as almost all UK residents are registered with a general practitioner and the population captured by CPRD practices is representative of the general population ^22^. Exposure is well measured as it is recorded from medical records and collected prospectively (avoiding recall bias). Our sampling methodology should have avoided any immortal time bias. We tried to reduce confounding by indication by only sampling cases and controls who could have been exposed to fibrates or glitazones because of a clinical indication. However, this design feature greatly reduced the number of cases and hence reduced our statistical power to detect modest effects. In addition, although we tried to adjust for diabetes severity using HbA1c, there was a considerable amount of missing data. We used multiple imputation to take this into account and believe this provides a less biased estimate than the complete case analysis. Adjustment for SES was only possible for linked practices which also had a negative impact on power but is unlikely to be biased as linkage was based on practice rather than individual consent. We could not test whether specific types of primary cancers such as lung cancer were more, less or equally likely to show a reduced risk.

There have been various *in vivo* studies that have suggested fibrate exposure may be protective for brain tumours, specifically gliomas, by modulating PPAR-α inhibition ^13,39-41^. There is similar evidence available for the consideration of glitazones as a treatment option from *in vivo* studies ^19,42-44^. However, prior to this study, there have been no other pharmacoepidemiological studies that examine whether fibrates or glitazones affect brain tumour risk and survival. A previously published study sought to investigate whether diabetes and use of anti-diabetic medications altered glioma risk in CPRD ^45^, however, the authors did not specifically look at glitazones as a drug of interest.

Our study has shown evidence that fibrates do not have an effect on glioma risk or prognosis. In contrast, longer exposure to glitazones was associated with a decreased risk of being diagnosed with primary or secondary brain tumours. Further research needs to try to replicate this finding using independent datasets preferably larger in size and/or with better data on glycaemic control and confounders. We hope our observations may lead to a better understanding of the pathophysiological mechanisms and potential therapies for the prevention of cancer. If the reduced risk with secondary tumours is causal and consistent across all types then this could be easily translated into a therapeutic option so that patients diagnosed with these cancers are started on these drugs as a tertiary prevention strategy to prevent brain metastases. This could be tested in a future clinical trial if these results are replicated in other cancer cohorts.

## Data Availability

Data were obtained from Clinical Practice Research Datalink under ISAC protocol number 18_149R. Researchers who wish to obtain the same data can do so under a license agreement, including the payment of appropriate license fees, between that third party and Clinical Practice Research Datalink.

